# Global, regional, and national burden of chronic kidney disease attributable to low physical activity from 1990 to 2021: an analysis of the Global Burden of Disease Study 2021

**DOI:** 10.1101/2025.02.08.25321935

**Authors:** Jinzhong Ji, Qing Guan, Yuning Ma, Gaozhan Ren, Mingxin Sun, Tingyang Huang, Weiqiang Lin, Xiuquan Lin, Hao Zhou

## Abstract

**Background:** Low Physical Activity (LPA) is a recognized risk factor for Chronic Kidney Disease (CKD). However, there is currently a lack of research reports addressing the global burden of CKD attributable to LPA.

**Methods:** We systematically described the burden of CKD attributable to LPA globally, regionally, and nationally using data from the Global Burden of Disease Study 2021. We examined the distribution by age, sex, and time trends. Furthermore, we conducted analyses on cross-national inequalities and frontier analysis. Additionally, we analyzed the distribution of CKD attributable to LPA burden among CKD subtypes.

**Results:** Between 1990 and 2021, the burden of CKD attributable to LPA significantly increased, reaching 40,918.47 deaths in 2021, with an age-standardized mortality rate (ASMR) of 0.50 per 100,000 population, 913,068.96 DALYs, and an age-standardized DALYs (ASDR) of 10.81 per 100,000 population. Over the next decade, the global burden of CKD attributable to LPA is projected to continue to rise, with an estimated ASMR of 0.90 per 100,000 population and an ASDR of 11.56 per 100,000 population by 2031. The 85-89 age group had the highest number of deaths, while the 70-74 age group had the highest DALYs, with both ASMR and ASDR increasing with age. Inequalities in CKD burden attributable to LPA exist across different Socio-Demographic Index (SDI) regions, with the Middle SDI region bearing the heaviest burden, but opportunities to alleviate CKD burden exist at all SDI levels. Globally, the highest proportion of CKD attributable to LPA was in hypertensive and diabetic nephropathy.

**Conclusions:** The burden of CKD attributable to LPA is increasing worldwide and is expected to continue rising over the next decade. Inequalities in CKD burden attributable to LPA exist. Globally, the burden of CKD attributable to LPA is primarily distributed among type 2 diabetes and hypertensive nephropathy. These findings underscore the importance of promoting physical activity in controlling CKD burden, especially targeting high-risk populations and regions.

## Introduction

Chronic Kidney Disease (CKD) represents a significant global public health challenge, with its prevalence on the rise and currently ranking as the seventh leading risk factor for mortality worldwide.^[1]^ As the global population ages, the burden of CKD and its risk factors are anticipated to escalate substantially. By 2050, the prevalence of CKD stages G3-G5 in certain regions may exceed 10%, resulting in considerable health and economic burdens, with particularly severe impacts on low-income countries.^[2]^ Mitigating the burden of CKD heavily relies on preventive measures addressing modifiable risk factors, such as lifestyle modifications including increased physical activity, healthy diet, and maintaining a healthy weight. Additionally, the implementation of cost-effective screening programs for early detection of high-risk individuals, and the widespread availability and affordability of novel nephroprotective medications are crucial to preventing CKD onset and slowing its progression.^[3-4]^

Low Physical Activity (LPA) is recognized as one of the risk factors for CKD, with a well-established association between physical inactivity and adverse outcomes in CKD patients. LPA not only increases the risk of cardiovascular disease mortality in CKD patients but also accelerates the decline in renal function^[5]^. Studies indicate a dose-response relationship between physical activity and CKD risk, estimating a 2% reduction in CKD risk for every 10 metabolic equivalents increase in physical activity.^[6]^ Therefore, LPA is an important and modifiable risk factor crucial for CKD prevention. It is noteworthy that the World Health Organization (WHO) has set a target to reduce physical inactivity by 15% from 2010 to 2030.^[7]^ However, the global burden of CKD attributable to LPA, as well as its temporal trends and age-sex patterns, remain inadequately explored. Accurate and up-to-date estimates of this burden are essential for planning research and developing evidence-based strategies for CKD prevention and management.

The 2021 Global Burden of Diseases Study (GBD 2021) systematically reviewed and integrated risk data from 88 different factors, including estimates of CKD burden attributable to low physical activity.^[8]^ This provides a unique opportunity to explore the epidemiology of CKD attributable to LPA on a global, regional, and national level. This paper aims to assess the temporal and spatial trends of CKD mortality and Disability-Adjusted Life Years (DALYs) attributable to LPA from 1990 to 2021, categorized by region, Socio-Demographic Index (SDI), country, sex, age group, and CKD type, to guide the allocation of physical activity resources and refine strategies for promoting physical activity to reduce the CKD burden, in response to the WHO ’ s 2030 physical inactivity target.

## Methods

### Definitions and Data Sources

CKD is clinically defined as a condition characterized by structural and functional abnormalities of the kidneys, persisting for a duration exceeding three months, which adversely impacts health.^[9]^ According to the GBD 2021, CKD is denoted by a permanent loss of kidney function, as indicated by the estimated glomerular filtration rate (eGFR) and the urine albumin-to-creatinine ratio (ACR). The corresponding International Classification of Diseases (ICD-10) codes for CKD include N18.1 through N18.9^[10]^. Physical activity levels are quantified in terms of total metabolic equivalent (MET) minutes per week, calculated by summing the frequency and duration of each activity along with the MET corresponding to the intensity of each activity. MET represents the ratio of working metabolic rate to resting metabolic rate, where one MET signifies the energy expenditure while sitting, equivalent to 1 kilocalorie per kilogram per hour^[11]^. The GBD 2021 assessed physical activity among adults aged 25 and older, focusing on activities lasting a minimum of ten minutes across all domains of life, including leisure/recreation, work, household, and transport^[11]^. Consistent with previous studies, GBD 2021 utilized two standardized questionnaires—the Global Physical Activity Questionnaire (GPAQ) and the International Physical Activity Questionnaire (IPAQ)—to collect self-reported data on activity intensity, duration, and frequency in a randomly sampled general adult population, subsequently quantifying physical activity through MET. Low physical activity is defined as engaging in less than 3,000 MET minutes of total physical activity per week^[11-12]^.

The deaths and DALYs of CKD attributable to LPA were estimated using a previously established comparative risk framework. In summary, the process began by determining the correlation between LPA and health outcomes, with the lowest observed exposure level taken as the theoretical minimum risk exposure level (TMREL) to estimate exposure levels and distributions. The overall attributable fraction (PAF) was computed using specialized formulas, incorporating variables such as age, sex, and year. Subsequently, the PAF was multiplied by the total disease burden to derive the specific disease burden attributable to LPA. All estimates are presented with 95% uncertainty intervals (UI), and the detailed modeling strategies and computational methods have been reported in The Lancet^[7]^

In this study, we obtained estimates for mortality, mortality rates, DALYs, and DALY rates attributable to LPA from the GBD 2021 website, including their 95% uncertainty intervals. Additionally, we utilized the SDI, a metric that quantifies the socio-demographic development of a region based on income, education, and fertility.

## Data Adjustment and Statistical Analysis

Firstly, an analysis of the age-standardized mortality (ASMR) and DALYs rate (ASDR) of CKD attributable to LPA was conducted to delineate its distribution across various global regions and countries. To further dissect the age and sex disparities in CKD attributable to LPA, the disease burden was quantified by stratifying the data by both sex and age. Additionally, a Spearman correlation analysis was employed to examine the relationship between the Sociodemographic Index (SDI) and the ASMR and ASDR, thereby elucidating the association between the burden of CKD attributable to LPA and SDI development.

The temporal trends of LPA attributable to CKD globally and across varying SDI regions from 1990 to 2021 were analyzed utilizing the Joinpoint regression model. The model facilitates the calculation of annual percentage changes (APC) and their accompanying 95% confidence intervals (CI) to describe trends within the specified time frame. Additionally, the average annual percentage change (AAPC) was calculated to assess overall trends from 1990 to 2021^[13]^. Furthermore, bayesian age-period-cohort (BAPC) models were also utilized to describe and forecast the burden of CKD attributable to LPA through 2031. This advanced model integrates historical data with probability distributions to estimate future patterns of CKD attributable to LPA, accounting for age, period, and cohort effects.^[14]^

This study assessed the cross-country inequality analysis of CKD burden attributable to LPA between different countries using the slope index of inequality (SII) and the concentration index (CI).^[15-16]^ To identify the maximum potential for CKD burden attributable to LPA for each country and region at given SDI levels, frontier analysis was conducted^[17]^. The detailed methodologies of of the cross-country inequality analysis and frontier analysis are described in the Supplementary Material.

In GBD 2021, CKD attributable to LPA includes CKD caused by type 2 diabetes mellitus (T2DM), glomerulonephritis, hypertension, and other or unspecified causes. We analyzed the distribution of CKD burden attributable to LPA by subtype across different regions, ages, sexes, and time periods to inform localized and personalized strategies to more effectively reduce CKD burden.

All statistical analyses and data visualizations were performed using R software (version 4.0.5), with P-values <0.05 considered statistically significant.

## Results

### Geographical Distribution of CKD Burden Attributable to LPA

In 2021, the global number of deaths from CKD attributable to LPA was estimated at 40,918.47 (95% UI: 16,172.11 to 72,562.93), with the ASMR of 0.50 per 100,000 (95% UI: 0.20 to 0.90). The percentage change in ASMR from 1990 to 2021 was 19% (95% UI: 3% to 32%) (Table S1). The global DALYs attributable to CKD attributable to LPA amounted to 913,068.96 (95% UI: 348,170.58 to 1,619,766.24), with the ASDR of 10.81 per 100,000 (95% UI: 4.14 to 19.18). The percentage change in DALYs rate from 1990 to 2021 was 12% (95% UI: 0% to 22%). Regionally, in 2021, the highest ASMR for CKD attributable to LPA was observed in Andean

Latin America at 1.06 per 100,000 (95% UI: 0.42 to 1.86), followed by Central Sub-Saharan Africa and North Africa and the Middle East (Table S1). In contrast, Eastern Europe had the lowest ASMR at 0.10 per 100,000 (95% UI: 0.03 to 0.19), followed by Central Asia and Central Europe. For ASDR, the highest rates were observed in Central Sub-Saharan Africa, followed by Andean Latin America and Central Latin America, whereas the lowest rates were in Eastern Europe, Central Asia, and Central Europe (Table S1).

At the national level, the top three countries with the highest ASMR in 2021 were American Samoa, Micronesia (Federated States of), and Marshall Islands, while the three countries with the lowest ASMR were Tajikistan, Ukraine, and Belarus (Figure 1). The highest ASDR were observed in American Samoa, Micronesia (Federated States of), and Marshall Islands, while the lowest rates were in Tajikistan, Ukraine, and Belarus (Figure 1).

**Figure 1.**
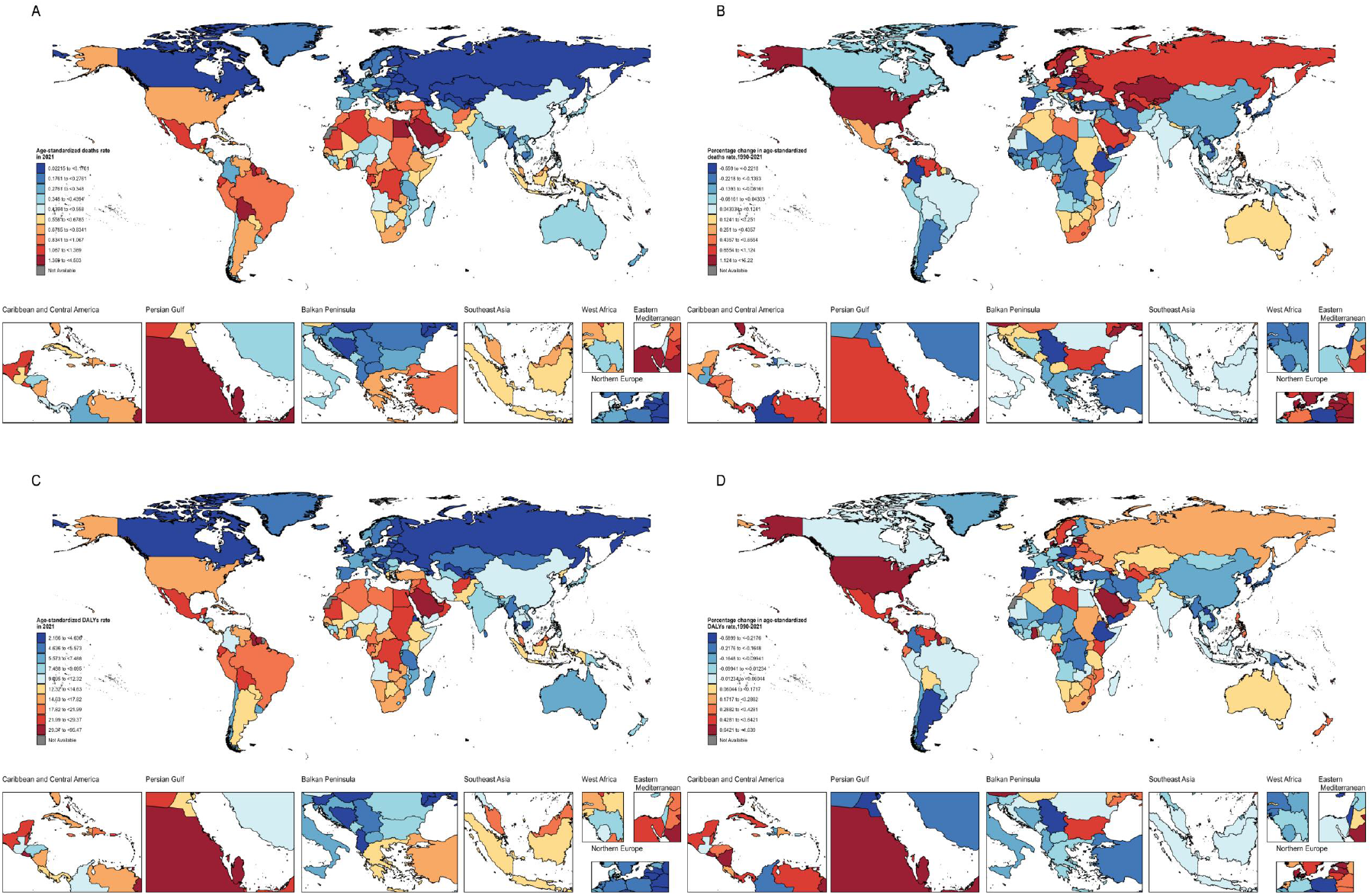
Geographical distribution of the burden of CKD attributable to LPA. ASMR (A)and ASDR (B) per 100,000 in 2021; Percentage change in ASMR (C) and ASDR (D) per 100,000 from 1990 to 2021.

### Age and Sex Patterns of CKD Burden Attributable to LPA

In 2021, the highest number of deaths from CKD attributable to LPA was in the 85-89 age group, totaling 6,112.22 (95% UI: 2,325.41 to 10,817.26), followed by the 80-84 age group with 6,020.97 (95% UI: 2,319.42 to 10,513.76) (Table S2). The mortality from CKD attributable to LPA increased with age, with the highest rates observed in those aged 85 and above (Fig 2A). The highest number of DALYs attributable to CKD attributable to LPA was in the 70-74 age group, with 119,205.98 (95% UI: 42,778.93 to 217,593.89) (Table S2). DALYs rates also followed a similar trend to mortality, increasing with age (Fig 2B, Table S3).

**Figure 2.**
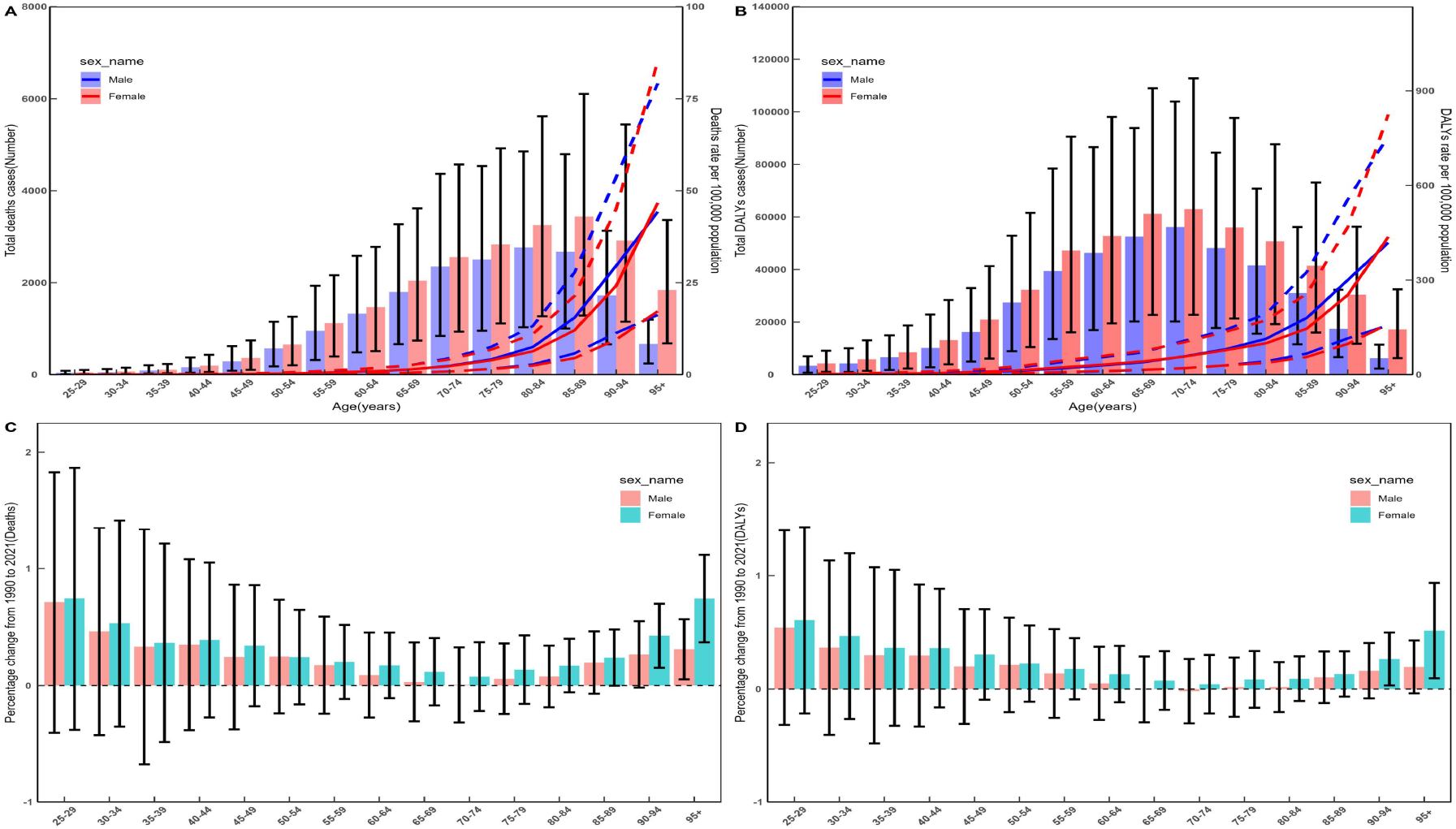
Age and sex patterns of CKD attributable to LPA burden. (A) Global CKD attributable to LPA death cases and ASMR per 100,000 in 2021; (B) Global CKD attributable to LPA DALYs and ASDR per 100,000 in 2021; Percentage change in ASMR (C) and ASDR (D) per 100,000 from 1990 to 2021.

In terms of sex distribution, both deaths and DALYs were higher in females compared to males. However, for mortality rates, males exhibited slightly higher rates and DALYs in the 75-94 age range, while females had higher rates in the 95+ age group. There were no significant differences in other age groups. Between 1990 and 2021, the age-standardized mortality rates and DALYs rates attributable to CKD attributable to LPA increased across all age groups, with the most significant increase observed in the 25-29 age group, followed by the 95+ age group (Fig 2C-D, Table S3).

### Temporal Trends in the Burden of CKD Attributable to LPA

Joinpoint regression analysis indicated that from 1990 to 2021, the global ASMR and ASDR for CKD attributable to LPA exhibited an overall increasing trend, with annual percentage changes (AAPC) of 0.58% and 0.37%, respectively. The most increase in ASMR occurred between 1998 and 2002 (APC = 1.92%), although a slight decline was noted from 2016 to 2021 (APC = -0.03%) (Figure S1A, Table S4). ASDR saw the most significant increase between 1997 and 2003 (APC = 1.24%), with a smaller increase from 2003 to 2021 (APC = 0.13%)(Figure S1B, Table S4).

The Bayesian Age-Period-Cohort (BAPC) model forecasts an overall increase in ASMR and ASDR for CKD attributable to LPA over the next decade, with ASMR projected to rise from 0.50 per 100,000 in 2021 to 0.51 per 100,000 by 2031. ASDR is expected to increase from 10.81 per 100,000 in 2021 to 11.68 per 100,000 by 2031 (Figure 3A-B, Table S5).

**Figure 3.**
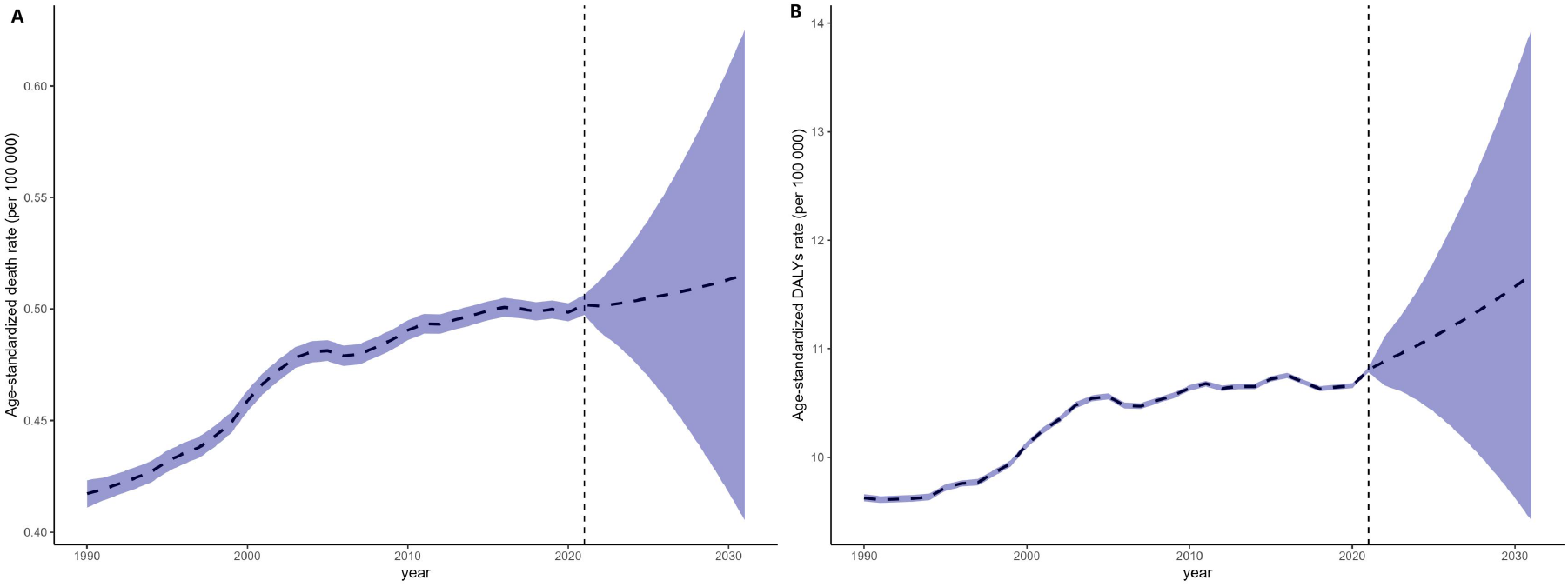
Trends in the burden of CKD attributable to LPA over the next 10 years. Prediction of global ASMR (A) and ASDR (B) per 100,000 of global CKD attributable to LPA.

### Burden Patterns of CKD Attributable to LPA by SDI

Among different SDI regions, the Middle SDI region had the highest ASMR at 0.59 per 100,000 (95% UI: 0.23 to 1.04), while the High-Middle SDI region had the lowest ASMR at 0.40 per 100,000 (95% UI: 0.16 to 0.71) (Table S1). Similarly, ASDR was highest in the Middle SDI region at 12.83 per 100,000 (95% UI: 4.81 to 22.67) and lowest in the High-Middle SDI region at 8.26 per 100,000 (95% UI: 3.23 to 14.47) (Table 1).

Globally, the observed ASMR and ASDR for CKD attributable to LPA were slightly below the expected values. In some regions, such as Central Sub-Saharan Africa, Andean Latin America, and Central Latin America had observed ASMR and ASDR higher than expected, whereas Eastern Europe, Central Asia, and Central Europe had observed values lower than expected. At the national level, ASMR and ASDR for CKD attributable to LPA showed a negative correlation with SDI, although the results were not statistically significant. Countries such as American Samoa, Micronesia (Federated States of), and Marshall Islands had observed ASMR and ASDR higher than expected, while Tajikistan, Niger, and Chad had observed rates lower than expected (Figure 4).

**Figure 4.**
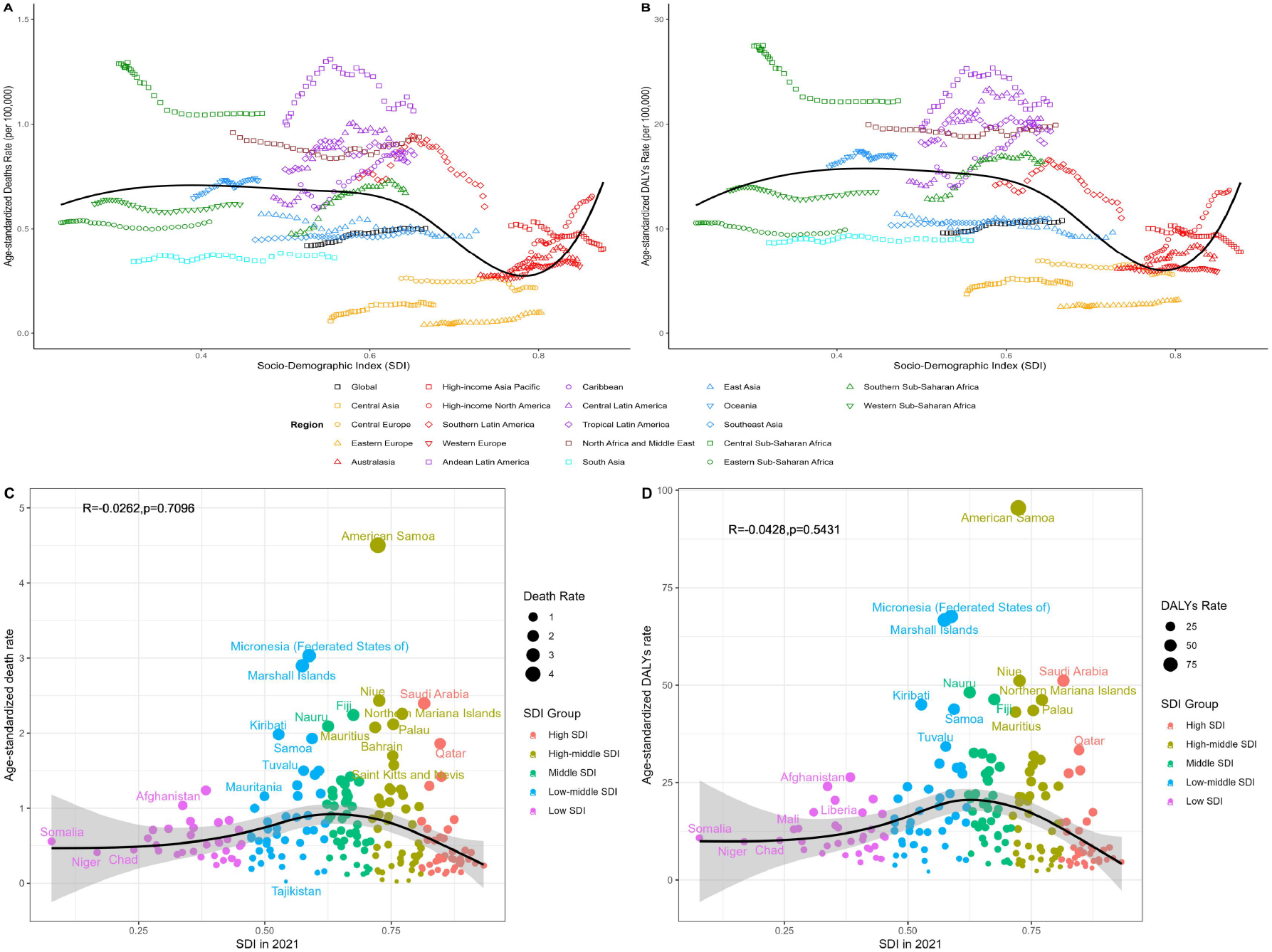
Association between CKD attributable to LPA burden and SDI. Association between ASMR (A) and ASDR (B) of CKD attributable to LPA and SDI by 21 GBD regional, association between ASMR (C) and ASDR (D) of CKD attributable to LPA and SDI by the national level.

### Cross-National Inequality Analysis of CKD Burden Attributable to LPA

In the burden of CKD attributable to LPA, both absolute and relative inequalities correlated with the SDI have been observed. Countries with lower SDI bear a disproportionately high burden. The inequality slope index indicates that the disparity in ASMR of CKD attributable to LPA between the highest and lowest SDI countries decreased from -0.21 in 1990 to -0.12 in 2021, while the disparity in ASDR decreased from -4.07 in 1990 to -2.74 in 2021 (Figure 5A-B, Table S6). This suggests a reduction in health inequality. Concentration curves for 1990 and 2021 show that the concentration curves for ASMR and ASDR of CKD attributable to LPA are predominantly above the equality line, indicating that the burden of CKD attributable to LPA is geographically concentrated in poorer regions (Figure 5C-D, Table S6).

**Figure 5.**
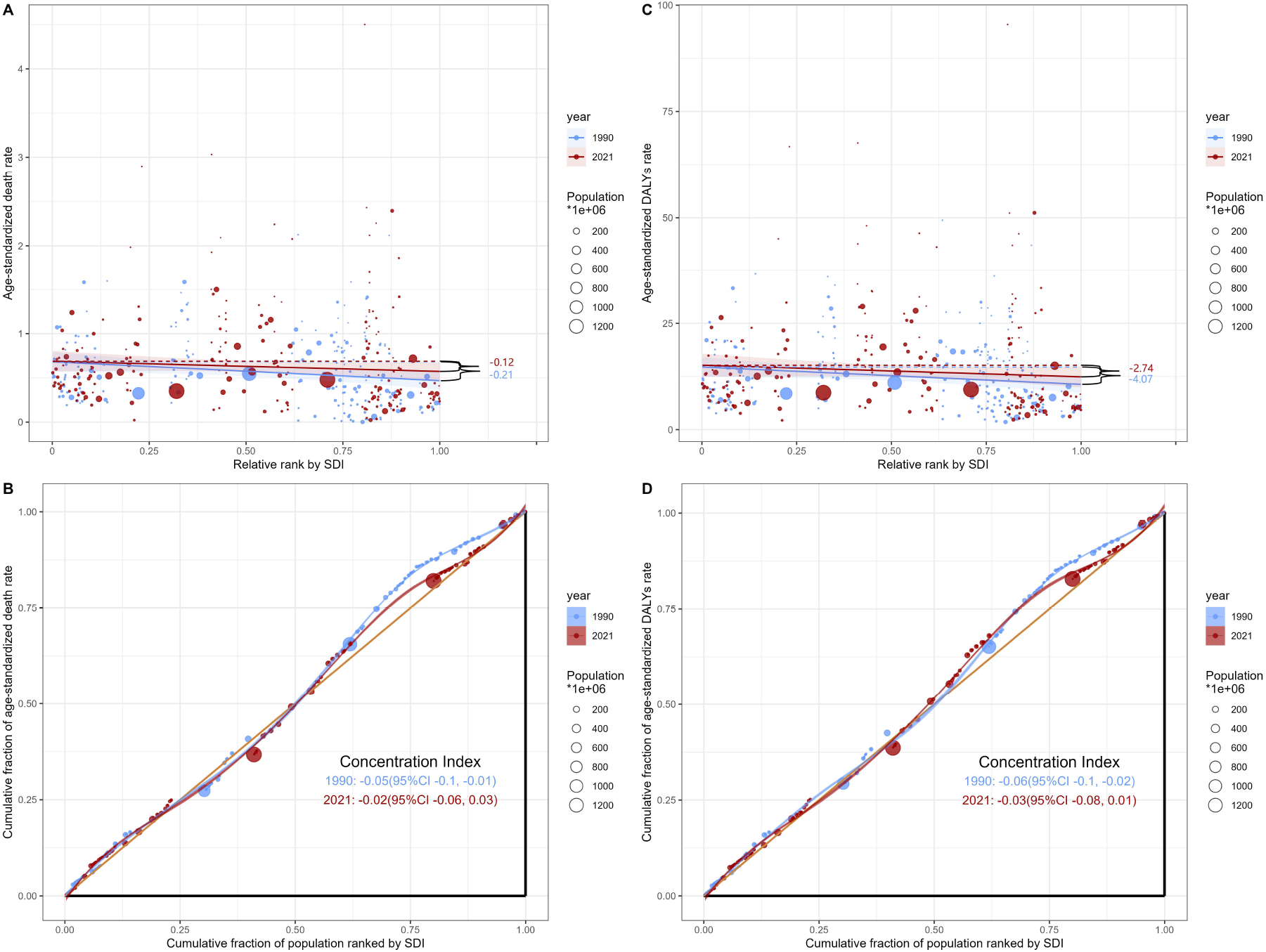
Cross-country inequality analysis of CKD attributable to LPA burden. Health inequality regression curve (A, C) and concentration curves (B, D) for ASMR and ASDR of CKD attributable to LPA in 1990 and 2021.

### Frontier Analysis of CKD Burden Attributable to LPA

Overall, the effective disparity for both ASMR and ASDR of CKD attributable to LPA has tended to increase from 1990 to 2021, given a constant SDI. Over time, the burden in most countries has increased (Figure 6). The greatest effective disparities are observed in the middle of the socio-demographic index spectrum (Figure 6A-B, Table S7). For ASMR, American Samoa, Micronesia (Federated States of), and the Marshall Islands are the countries most likely to narrow the gap. Among countries with lower SDI, Somalia, Niger, and Eritrea have shown excellent performance with minimal effective disparities. Conversely, some higher SDI countries such as the United States of America, Denmark, and Australia have exhibited larger effective disparities. The distribution trend of ASDR is broadly consistent with that of ASMR.(Figure 6C-D, Table S7).

**Figure 6.**
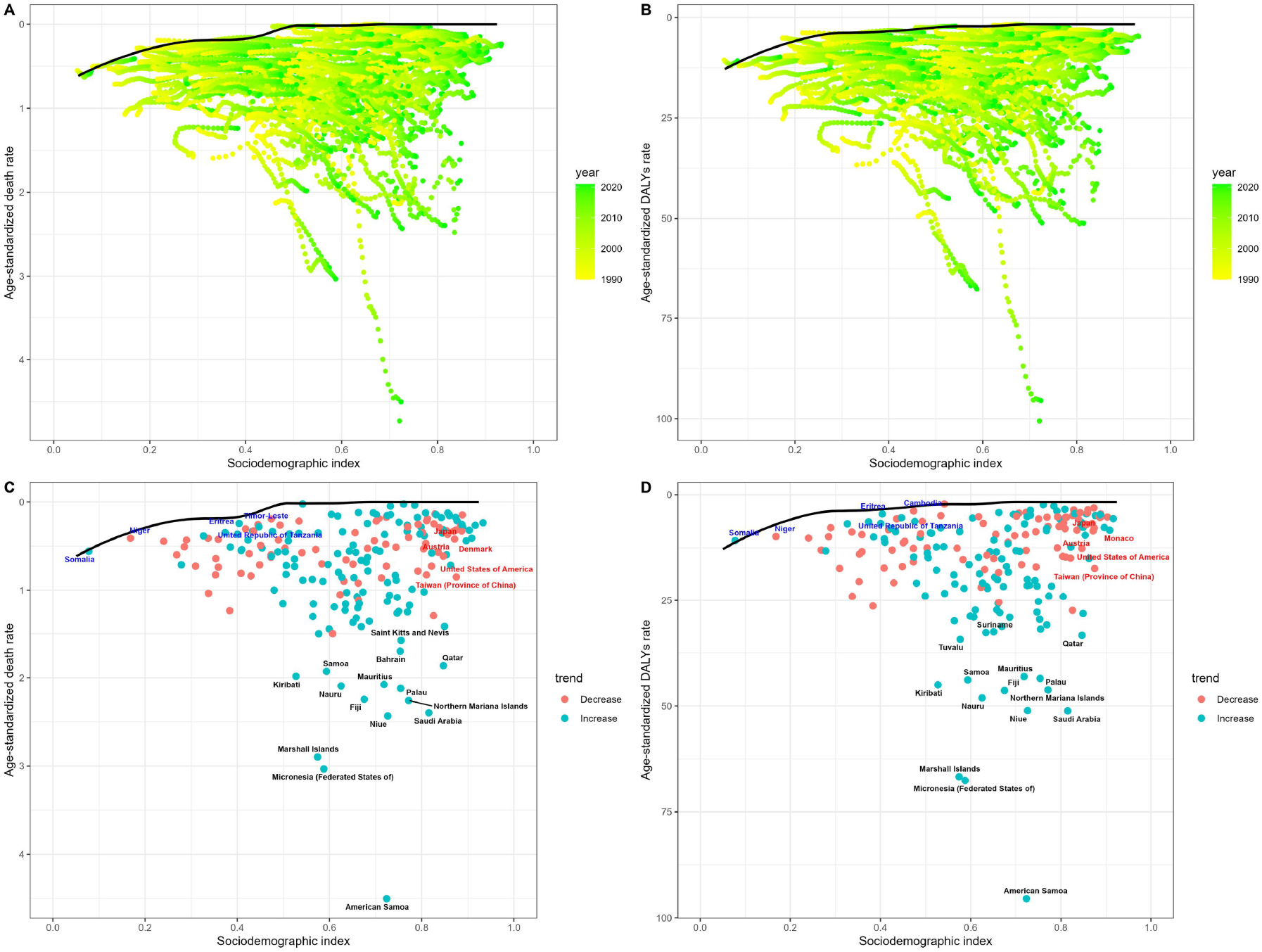
Frontier analysis based on the SDI and the burden of CKD attributable to LPA. Frontier analysis in the ASMR (A) and ASDR (B) of CKD attributable to LPA from 1990 to 2021, and the ASMR (C) and ASDR (D) of CKD attributable to LPA in 2021. Note: The top 15 countries with the greatest effective disparities are marked in black. Regions with high SDI values (>0.85) and substantial effective disparities are highlighted in red, while those with low SDI values (<0.5) and smaller effective disparities are indicated in blue. The boundary, depicted in pure black, represents the potential ASMR and ASDR achievable based on SDI. Actual values for countries and regions are denoted by dots, with increases in the burden of CKD attributable to LPA from 1990 to 2021 represented by blue dots and decreases by red dots.

### Distribution of CKD Burden Attributable to LPA by Etiology

In 2021, among different causes of CKD, T2DM-related CKD was the leading type of CKD burden attributable to LPA globally, followed by hypertension-related CKD. Similar results were observed across different SDI regions, with T2DM and hypertension-related CKD being the major contributors to CKD burden attributable to LPA (Figure S2A-B). Across different age groups, T2DM-related CKD is the principal type of CKD burden attributable to LPA for both males and females (Figure S2C-D). However, for ASMR, hypertension-related CKD ranks second across all age groups, while for ASDR, other causes of CKD rank second among males, and hypertension-related CKD ranks second among females. From 1990 to 2021, the global ASMR for CKD attributable to LPA was predominantly attributed to type 2 diabetes-related CKD, followed by hypertension-related CKD. Similarly, over the past three decades, the global ASDR for CKD attributable to LPA has also been primarily attributable to type 2 diabetes-related CKD, followed by other causes of CKD (Figure S2E-F).

## Discussion

The association between LPA and CKD has been well-established. Regular physical exercise, ranging from light walking to vigorous activity, has demonstrated significant health benefits.^[18]^ Reports indicate that individuals engaging in the highest levels of physical activity exhibit a lower risk of CKD compared to those who are inactive or engage in minimal activity.^[19]^ Previous studies have shown that exercise benefits adults with CKD, renal failure, or kidney transplantation by reducing chronic inflammation, enhancing cardiopulmonary function, and improving muscle and bone strength as well as metabolic markers.^[20]^ Current guidelines recommend that non-dialysis CKD patients aim for a cumulative 150 minutes of moderate-intensity aerobic exercise per week, gradually increasing from their current levels. Dialysis CKD patients should engage in 150 minutes of moderate-intensity activity (or 75 minutes of vigorous activity) per week or a combination of both^[7]^. Various interventions, such as media campaigns, social support for physical activity in communities and workplaces, and improved access to exercise facilities, can help increase physical activity.^[21]^ Therefore, LPA is a modifiable risk factor for CKD, highlighting the urgent need for more comprehensive health interventions and public awareness campaigns to address this issue.

The burden of CKD attributable to LPA varies globally. Our results indicate that, in 2021, the burden of CKD attributable to LPA was highest in middle SDI regions, followed by low-middle and low SDI regions, with high-middle and high SDI regions experiencing lower burdens. This phenomenon may be attributed to higher socio-economic status individuals engaging in more leisure-time physical activity compared to those with lower socio-economic status.^[22]^ Additionally, as industrialization and economic development progress, people in lower SDI regions are likely to engage in jobs that demand less physical activity attributable to changes in transportation, entertainment, and cultural values.^[23]^ Notably, from 1990 to 2021, the burden of CKD attributable to LPA increased in all SDI regions except low SDI regions, with particularly rapid growth in high SDI regions. Reports indicate that in high-income regions of Western Europe and North America, physical activity levels are significantly lower than the global average in higher socio-economic areas, which may contribute to the increased burden in these regions.^[24]^ However, this trend differs from previous findings. The GBD 2019 report on LPA burden revealed a substantial decrease in ASDR in high SDI regions from 1990 to 2019, whereas other regions saw increases during the same period^[10]^. This indicates a disparity between CKD burden attributable to LPA and the overall disease burden attributable to LPA. Frontier analyses indicate disparities among countries with varying SDIs, particularly in middle SDI countries, yet opportunities to alleviate CKD burden exist at all SDI levels. Therefore, efforts to promote physical activity should be intensified in both developed and developing nations.

Our study demonstrates that the ASMR and ASDR for CKD attributable to LPA increase with age, with the elderly bearing a heavier burden of CKD attributable to LPA compared to younger individuals. This pattern is consistent with the distribution of the overall disease burden attributable to LPA^[10]^. Reports show that physical activity declines sharply with age.^[25]^ Surprisingly, despite the relatively low burden of CKD attributable to LPA in the 25-29 age group, this group has experienced the fastest growth in ASMR and ASDR from 1990 to 2021. This trend suggests that insufficient physical activity among younger individuals is a growing concern and warrants attention to prevent worsening disease burden among youth. Recent reports emphasize the importance of implementing physical activity interventions at a relatively young age to prevent, reduce, or delay functional decline in later years.^[26]^ Thus, while it is crucial to focus on physical activity for the elderly, engaging in early and active exercise in individuals before organ damage occurs may also be beneficial in preventing CKD burden in older populations.

Our findings indicate that the burden of CKD attributable to LPA primarily affects T2DM and hypertension-related CKD. It is well established that diabetic nephropathy and hypertensive nephropathy are the most common subtypes of CKD, and LPA is also a risk factor for T2DM and cardiovascular diseases, with the burden of these conditions continuously increasing.^[27]^ Research shows that low to moderate-intensity aerobic exercise has nephroprotective effects in T2DM patients, enhancing insulin sensitivity, reducing microalbuminuria, and improving lipid levels.^[28]^ Another study focusing on diabetic patients revealed that moderate activity is associated with significantly lower risks of adverse kidney outcomes and reduced incidence of new-onset microalbuminuria compared to lower levels of exercise.^[29]^ A meta-analysis of 29 studies assessing the quantitative dose-response relationship between physical activity and hypertension found that every 10 metabolic equivalent hours per week reduction in leisure-time physical activity is associated with a 6% increase in the risk of developing hypertension.^[30]^ Therefore, enhancing physical activity in CKD patients, especially those with T2DM and hypertension-related nephropathy, should be prioritized.

This study has several limitations. Firstly, it is subject to the common constraints inherent in all GBD studies, particularly with respect to the heterogeneity of data collection methods and sources, as well as the quality and completeness of data. Secondly, due to data limitations, our analysis focused solely on the people aged 25 and above. Finally, it is important to note that not all countries and years have related data on low physical activity, especially in low-development countries. The GBD project employs statistical methods to estimate the disease burden associated with low physical activity in regions with sparse data, providing estimates with appropriate uncertainty rather than omitting critical information.

In summary, the burden of CKD attributable to LPA has increased globally and is expected to continue rising over the next decade. Specifically, there are health inequalities in the burden of CKD attributable to LPA, with the highest burden observed in middle SDI regions and among the elderly. Additionally, the global burden of CKD attributable to LPA is primarily associated with T2DM and hypertension-related nephropathy. These findings highlight the necessity for tailored exercise prescriptions for specific populations. Furthermore, it is crucial to raise public awareness regarding the importance of physical activity in managing CKD burden, as well as to encourage policymakers to develop strategies for implementing and equitably distributing resources for physical activity.

## Data Availability

All data produced in the present study are available upon reasonable request to the authors

https://ghdx.healthdata.org/gbd-2021/results

## Author Contributors

HZ and XQL conceived and designed the study. JZJ, YNM and QG drafted the manuscript. JJZ, QG, YNM, GZR, MXS and TYH analyzed the GBD data. HZ XQL and WQL were responsible for the article checking and reviewing. JJZ, GQ, HZ and XQL accessed and verified the data. All authors revised the report and approved the final version before submission. HZ had access to all the data in the study and had final responsibility for the decision to submit for publication.

## Disclosures

All authors declare no conflict of interest.

## Funding

This work was supported by the Huadong Medicine Joint Funds of the Zhejiang Provincial Natural Science Foundation of China [No. LHDMZ24H050001], National Natural Science Foundation of China [No. 82074166], Fujian Provincial Natural Science Foundation [No. 2018J01121], and Fujian Provincial Health Technology Project [No.2020GGA026].

## Acknowledgments

We highly appreciate the work by the GBD 2021 collaborators.

## Data Sharing Statement

The data of this study are publicly available at the GBD 2021 website (https://ghdx.healthdata.org/gbd-2021/results). All data also will be made available on request to the corresponding author.

## Notes

### Competing Interest Statement

The authors have declared no competing interest.

### Author Declarations

Ethical approval and consent were not required for this study, as it utilized publicly available data that were aggregated and anonymized.

